# CD56^bright^CD16^-^ NATURAL KILLER CELLS ARE IN HIGHER COUNTS IN THE UMBILICAL CORD BLOOD THAN IN THE PERIPHERAL BLOOD

**DOI:** 10.1101/2021.06.09.21258083

**Authors:** Vinicius Campos de Molla, Míriam Cristina Rodrigues Barbosa, Alfredo Mendrone, Matheus Vescovi Gonçalves, Eliza Kimura, Fabio Guirao, Mihoko Yamamoto, Celso Arrais-Rodrigues

## Abstract

Umbilical cord blood (UCB) is an alternative source for hematopoietic stem cells allogeneic hematopoietic stem cell transplantation in the absence of compatible donor. UCB transplantation has a lower incidence of chronic graft versus host disease (GvHD) but is associated with slower engraftment and slower immune reconstitution as compared to other sources. Dendritic cells (DC) and Natural Killer cells (NK) play a central role in the development of GVHD, the graft versus leukemia (GvL) effect, and in the control of infectious complications. We quantified by multiparametric flow cytometry monocytes, lymphocytes, NK cells, and DC, including their subsets, in UCB samples from 54 healthy newborns and peripheral blood (PB) from 25 healthy adult volunteers. In the UCB samples, there were higher counts of CD56brightCD16-NK cells (median 0.024×109/L), as compared to the PB samples (0.012×109/L, P<0.0001), CD56dimCD16bright NK cells (median 0.446×109/L vs. 0.259×109/L for PB samples, P= 0.001), and plasmacytoid dendritic cells (pDC, median 0.008×109/L for UCB samples vs. 0.006×109/L for PB samples, P= 0.03). Moreover, non-classical monocytes counts were lower in UCB than in PB (median 0.024×109/L vs. 0.051 x109/L, respectively, P< 0.0001). In conclusion, there were higher counts of NK cells and pDC, and lower counts of non-classical monocytes in UCB than in PB from healthy individuals. These findings might explain the lower incidence and severity of chronic GVHD although maintaining the GVL effect in UCB transplants recipients as compared to other stem cell sources.

**Highlights:**

- CD56brightCD16- NK cells are more frequent in UCB than in PB.
- Plasmacytoid dendritic cells are more frequent in UCB than in PB.
- Non-classical monocytes are more frequent in PB than in UCB.

## INTRODUCTION

In the last few years, allogeneic hematopoietic stem cell transplantation (HSCT) has improved clinical outcomes, with less transplant-related mortality (TRM) and better overall survival (OS) (1). Despite the advance in the knowledge of immune reconstitution in HSCT, there are still gaps in this field, such as the mechanisms of immunotolerance, graft-versus-host-disease (GvHD) or graft-versus-leukemia (GvL) effect (2).

Umbilical cord blood (UCB) transplant was performed for the first time in 1988 (3). UCB transplant is associated with delayed engraftment, increased risk of infections, greater engraftment failure, and worse TRM, as compared to bone marrow (BM) and peripheral blood (PB) sources (4). However, UCB transplant usually causes less acute GvHD (aGvHD) and chronic GvHD (cGvHD) with similar rates of leukemia-free survival (5). This could be explained by distinctive patterns of immune reconstitution according to graft source (6), regardless of the conditioning regimen (7).

Dendritic cells (DC) are antigen-presenting cells, making the connection between innate and adaptive immunity (8), and also coordinating the immune system through activation and stimulation of T and B cells, tolerance by removal of self-reactive T cells (9) and linkage with regulatory T cells (10). So, DC play a key role in immune system control. In the PB, DC are present in two subsets: myeloid (or conventional) DC (mDC) and plasmacytoid DC (pDC). The pDC (CD123+CD11c) produce interferon (IFN) I and are implicated in viral immune response, immune tolerance, and memory. At the same time, mDC (CD123-CD11c+) are responsible for a proinflammatory effect. Each DC subset is flexible in vivo and its responses vary according to factors such as their activation state, the nature of the stimulus received, and the inflammatory microenvironment (11). In the immune recovery after HSCT, DC play a relevant role in aGvHD, cGvHD, GVL and host response to infections (12), and a poor DC reconstitution is associated with increased risk of relapse and poor survival (13,14).

NKT cells (NKT) are T cells that express NK receptors, including NK 1.1 (CD161c) and semi-invariant CD1d-restricted αβ TCRs. There are two types of NKT: type I NKT (or iNKT) that express Vα24-Jα18, and type II NKT that express other TCR different than Vα24-Jα18 (15).

Natural killer cells (NK) are components of the innate immune system, eliminating tumor and infected cells, promoting cellular lysis in the absence of the HLA class I receptor. NK cells are categorized as CD56dimCD16bright (90% of NK in PB) with predominant cytotoxic effect through the release of granzyme B and perforin, and CD56brightCD16-related to INF-γ and tumor necrosis factor α (TNF-α) secretion (16). NK are the first cells to recover after HSCT and strongly contribute to the GvL effect, possibly due to killer-cell immunoglobulin-like receptor (KIR) mismatch (17). It has been shown that low counts of NK CD56brightCD16-after HSCT are associated with a lower survival rate (18), and KIR mismatches improve survival (19).

To better understand the mechanisms involved in different outcomes in HSCT using UCB or PB, we compared the composition of immune-related cells in these two HSC sources (monocytes, B lymphocytes, T cells, NK cells, and DC and their subsets) in samples from UCB and PB.

## MATERIALS AND METHODS

### Population and design

This was an analytical case-control study. Samples from 54 UCB and 25 PB were collected between September 2015 and July 2017. UCB samples from healthy newborns, with a minimum gestational age of 35 weeks, were collected at the Cellular Therapy Center of Hospital Sírio Libanês (before freezing). UCB from neonates with any antenatal risk factors were excluded. PB samples were collected from healthy adult blood donors at Hospital São Paulo / UNIFESP Blood Center. Blood donors and pregnant women who agreed to donate UCB (both ≥ 18 years old), without any documented chronic illness or infectious disease entered the study. Blood donor volunteers with blood transfusion in the last 3 months were excluded.

The study was approved by the local ethics committee of the participating centers and all volunteers (blood donors and mothers) gave their informed consent before entering the study, in accordance with the Declaration of Helsinki.

### Cells Identification and count by Flow Cytometry

Fresh EDTA-anticoagulated PB or UCB samples were processed in up to 24 hours after collection. Total nucleated cells from UCB were quantified by Coulter AcT Diff 2® (Beckman Coulter, Brea, EUA) and PB cells by Cell Dyn Ruby® (Abbott, Illinois, EUA) and erythroblasts were quantified by microscopy to correct the leukocytes count. Leukocytes and subsets were analyzed by flow cytometry.

Cells were stained using an 8-color monoclonal antibodies panel: CD16 FITC / CD56 & CD4 PE / CD11c PerCP Cy5.5 / CD8 & CD19 PC7 / CD123 APC / CD3 & CD14 APC-H7,/ HLA-DR Pac Blue/ CD45 OC-515, by stain-lyse-wash method. The reagents were purchased from: Cytognos, Salamanca (CD16, CD56, CD4, CD45), BDB, San Jose CA(CD11c, CD3, CD14), Immunotech Brea (CD8), Immunotech Marseille (CD19), Biolegend, San Diego (HLA-DR, CD123), Data acquisition (500×103 total events) was performed using FACSCANTOII flow cytometer (BDB-San Jose, CA) and FACsDIVA® software (BDB-San Jose, CA) and Infinicity® software (Cytognos, SL) was used for data analysis. NK cells (CD3-, CD19-, CD14-, and CD56+), were classified as CD56brightCD16- or CD56dimCD16bright. Monocytes (CD45+, CD11c+, and HLA-DR+) were classified as: classic (CD14++CD16-), intermediate (CD14++CD16+) and non-classical (CD14dimCD16++). Dendritic cells (HLA-DR++, CD3-, CD19-, CD14-, and CD56-) were classified as pDC (CD123+/ CD11c-), and mDC (CD123-/ CD11c++).

### Statistical Methods

Descriptive statistical analysis was reported by using percentages (categorical variables) and ranges and medians (continuous variables). Comparisons between UCB and PB were performed using the Mann-Whitney test. A P value of <.05 was considered significant. SPSS version 21.0 (SPSS Inc., Chicago, IL) was used for all statistical analyses.

## RESULTS

### Clinical Characteristics

Seventy-nine samples (54 UCB and 25 PB) were obtained. UCB samples were collected from newborns with a median gestational age of 40 weeks (range: 36 to 43 weeks), and birth weight of 3.263kg (range: 2.530 to 4.005kg). Twenty-nine (54%) newborns were male. Vaginal delivery was more commonly used (74%) than the cesarean section (26%). PB samples donors median age was 33 years old (range 18 to 66 years old), and 15 (60%) were male.

### Absolute counts of WBCs and subpopulations

Total white blood cell (WBC) count was higher in UCB than in PB (10.782×109cells/L vs. 6.980×109cells/L, P<0.0001). The total number of cells from most lineages (neutrophils, eosinophils, monocytes, B cells) were higher in UCB, while total T cells, NK cells, and total DCs were similar in both groups (Table 1). Among subpopulations, many differences were noted: non-classical monocytes, double positive CD4+CD8+ T cells and double negative CD4-CD8-T cells were in higher counts in PB than in UCB (0.051×109cells/L vs. 0.024×109cells/L, P<0.0001; 0.005×109cells/L vs. 0.001×109cells/L, P<0.0001; 0.054×109cells/L vs 0.039×109cells/L, P=0.005, respectively). However, pDC were in lower counts in PB than in UCB (0.006×109cells/L vs. 0.008×109cells/L, P=0.03, Table 2). No significant statistical difference was observed in mDC counts between the groups.

### Relative frequencies of WBCs and subpopulations

The proportion of B cells and NK cells were higher in UCB samples when compared to PB samples (16% vs. 11%, P<0.0001; 20% vs. 11%, P<0.006, respectively).

In terms of relative frequencies of subpopulations, classical monocytes were the predominant subset being present in a higher count in UCB vs PB (89% vs 77%, P<0.006×109, respectively. Classical monocytes: non-classical monocytes ratio was 28.3 in UCB and 7.0 in PB (P<0.0001).

T cells CD4+ were more prevalent than T cells CD8+ in both PB and UCB (CD4: CD8 ratio 1.8 and 2.3, respectively), while T cells CD8+ were more frequent in PB (31%), rather than UCB (27%, P=0.03).

NK cells CD56dimCD16bright counts were the predominant subset in both samples (96% and 93%, PB and UCB, respectively). The ratio between the subtypes (NK cell CD56dimCD16bright:CD56brightCD16-) was lower in UCB (13.7) vs. PB (24, P=0.03). CD56dimCD16bright proportions of pDC in UCB as compared to PB from normal adults

The mDC were the predominant subtype in both PB and UCB, being 3 times the number of pDC in PB and 1.8 times the number of pDC in UCB. However, there was no difference in mDC absolute counts between the two groups. The pDC proportion was 25% in PB and 36% in UCB (P<0.0001). The mDC:pDC ratio was 3.0 in PB vs.1.8 in UCB (P<0.0001).

The CD4:CD8 ratio was higher in UCB (2.3) than in PB (1.8, P=0.01), and the NK cell CD56dimCD16bright:CD56brightCD16-ratio was lower in UCB (13.7) vs. PB (24, P=0.03). Intermediate monocytes: non-classical monocytes ratio was 28.3 in UCB and 7.0 in PB (P<0.0001). The relative distribution of WBCs and subpopulations are listed in Table 1 and Table 2.

## DISCUSSION

In the present study, we compared concentrations (cells per liter) and frequencies (%) of WBCs and subsets in UCB and PB from healthy individuals and observed higher counts and frequencies of CD56brightCD16-NK cells and pDC, and lower counts and frequencies of non-classical monocytes in the UCB. To the best of your knowledge, this is the first study that compared subsets of NK (CD56dimCD16bright, and CD56brightCD16-) between UCB and PB. NK CD56brightCD16-is a more immature NK cell, presenting a more humoral immune response profile, secreting INF-γ, TNF-α, IL-10, IL-13, and granulocyte-macrophage colony-stimulating factor, rather than the direct cytotoxic effect of the NK CD56dimCD16bright. They usually do not present KIR receptors and present higher expressions of the heterodimer CD94/NKG2A (major inhibitor receptor), which confers a lower alloreactivity than their more mature CD56dimCD16bright counterpart (20). Therefore, higher counts and higher frequencies of CD56brightCD16-NK cells and plasmacytoid dendritic cells in UCB as compared to PB could possibly lead to a more tolerogenic profile against host cells in HSCT recipients.

Of interest, monocyte maturation was clearly more evident in PB than in UCB samples. PB showed high proportions of the late phases of monocyte differentiation, like intermediate and non-classical monocytes, which was also observed by some previous studies (21). These populations represent a shift towards higher antigen-presenting activity, and they are considered as monocyte-derived DCs (22), and they are being also related to autoimmune diseases, such as rheumatoid arthritis and Crohn’s disease (23).

We found higher counts of pDC in UCB than in PB, corroborating the findings of Prabhu SB et al (21); pDC produces IFN type I, which is related to antiviral response, immune tolerance, and anti-tumor immunity through secretion of type-I IFN and TNF- α (24). Moreover, our group previously demonstrated that grafts with high pDC content lead to lower aGVHD and lower mortality risk after HSCT (18).

GvHD is a reaction caused in part by donor alloreactive T CD8+ and T cell depletion (*in vivo* or *ex vivo*) is a common strategy to mitigate this complication (25,26). Here we demonstrated a higher concentration of T CD8+ in PB when compared with UCB, as has been shown in previous studies (27–29).

All these findings in PB: more mature monocytes, a higher concentration of T CD8+, and lower counts of pDC and CD56brightCD16-NK as compared to UCB might explain the higher incidence of cGvHD when PB is used as stem cell source for HSCT (30). Conversely, the more tolerogenic profile observed in UCB as compared to PB could explain the lower risk of cGvHD with similar relapse rates, as evidence of the sustained GvL effect (5,31,32).

Our study has limitations, such as small sample size, and the use of PB samples without the stimulation of granulocyte colony-stimulating factors, as commonly used for HSCT, which could change the cells counts and frequencies.

**Figure 1.**
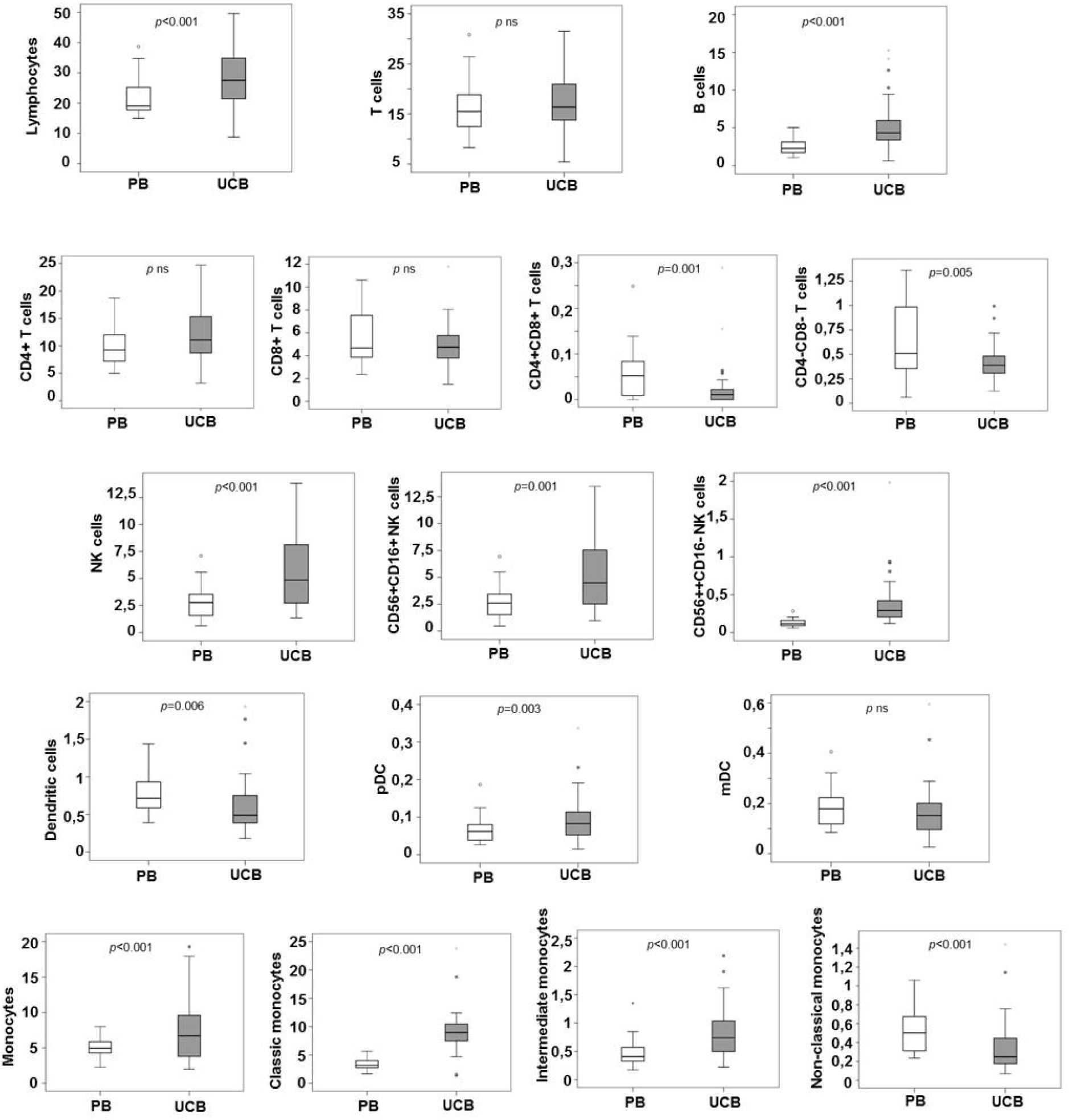
Concentrations of lymphocytes, NK cells, dendritic cells, monocytes and respective subsets in PB and UCB samples.

In conclusion, we observed higher counts of CD56brightCD16-NK cells and pDC and lower counts of non-classical monocytes in UCB as compared to PB from healthy individuals. As we propose in Figure 2, our findings demonstrate that UCB cells profile is more tolerogenic than that of PB and this finding might explain the lower incidence and severity of cGVHD in UCB recipients as compared to other stem cell sources, although maintaining the GVL effect. Future research is needed for a better understanding of immune reconstitution in HSCT, and to explain the differences regarding immune reconstitution, GvHD risk, GvL effect and other clinical outcomes in the HSCT among different stem cell sources.

**Figure 2.**
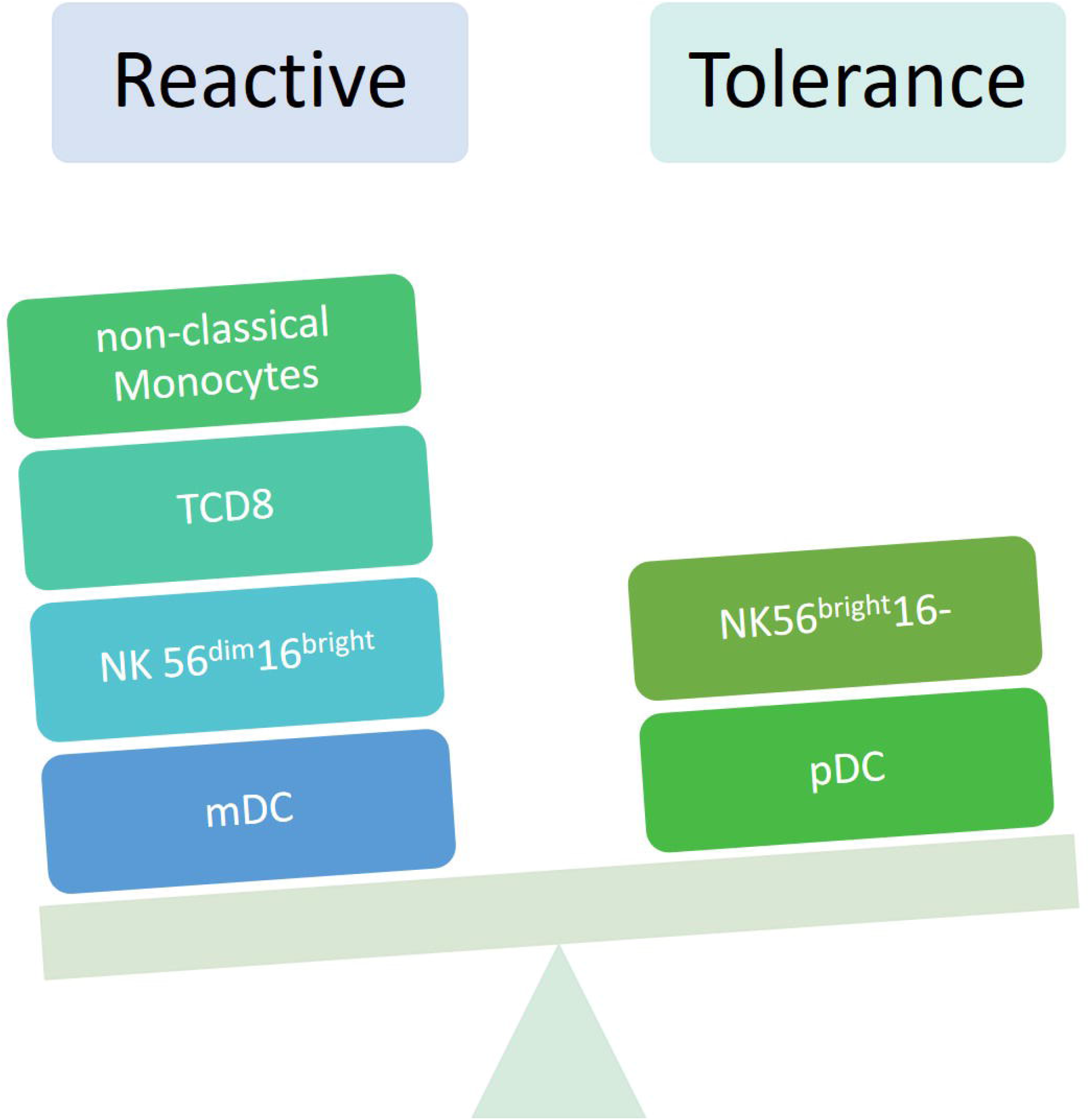
Differences in the profile of cell tolerance and reactivity: the profile of UCB cells is more tolerogenic than that of PB. Abbreviations: *mDC* myeloid dendritic cells, *pDC* plasmacytoid dendritic cells, *NK 56*^*dim*^*16*^*bright*^ natural killer cells CD56^dim^CD16^bright^, *TCD8* T cells CD8+, *NK 56*^*bright*^*16+* natural killer cells CD56^bright^CD16-.

## Data Availability

The data is available in our database.

## ACKNOWLEDGMENTS

M.C.R.B. was supported by Coordenação de Aperfeiçoamento de Pessoal de Nível Superior (CAPES, no. 001). V.C.M. was supported by Conselho Nacional de Desenvolvimento Científico e Tecnológico (CNPq process no. 141575/2018-2).

Authorship statement: M.C.R.B., V.C.M, M.V.G., and C.A.R. designed research, performed research, analyzed data, and wrote the paper; V.C.M, M.V.G., and C.A.R. performed statistical analysis; A. M. J. and C. A. A provided cord blood samples, M.C.R.B., M.Y. and M.V.G. performed flow cytometry analysis, critical review, and revised the manuscript. All authors drafted and approved the manuscript and agreed with its submission.

